# Preclinical Basal Ganglia Neurodegeneration in Female X-Linked Dystonia-Parkinsonism Carriers

**DOI:** 10.64898/2026.09.14.26362974

**Authors:** Henrike Hanssen, Cid Czarina E. Diesta, Marcus Heldmann, Jackson Dy, Jeffrey Tantianpact, Jan Uter, Max Brand, Björn-Hergen Laabs, Raymond L. Rosales, Ana Westenberger, Christine Klein, Jean Q. Oropilla, Norbert Brüggemann

**Affiliations:** Section for Movement Disorders, Department of Neurology, University of Luebeck and University Medical Center Schleswig-Holstein, Campus Luebeck, 23562 Luebeck, Germany; Institute of Neurogenetics, University of Luebeck, 23562 Luebeck, Germany; Center of Brain, Behavior and Metabolism, University of Luebeck, 23562 Luebeck, Germany; Makati Medical Center, Makati City, 1229 Metro Manila, Philippines; Asian Hospital and Medical Center, 1780 Metro Manila, Philippines; Institute of Medical Biometry and Statistics, University of Luebeck, Luebeck, Germany; University Medical Center Goettingen, Goettingen, Germany; Department of Neurology and Psychiatry, University of Santo Thomas, 1015 Metro Manila, Philippines

## Abstract

**Background and Objectives:** X-linked dystonia-parkinsonism (XDP) is a severe neurodegenerative movement disorder caused by a retrotransposon insertion with a polymorphic hexanucleotide repeat expansion in the *TAF1* gene. While the disease predominantly affects males, female carriers may exhibit a variable phenotypic expression. Neurobiological alterations in asymptomatic female mutation carriers (fMC) remain largely unexplored. We aimed to investigate whether fMC exhibit neurodegenerative changes despite the absence of overt clinical manifestations and to assess their clinical, genetic, and longitudinal correlates using multimodal MRI.

**Methods:** In this cross-sectional and longitudinal multimodal study, 103 female relatives of XDP patients underwent clinical, genetic, and MRI assessments. Forty-two were identified as fMC and 61 as healthy controls (HC). Structural T1-weighted MRI was analyzed using voxel-based morphometry and subcortical volumetry, while susceptibility-weighted imaging assessed iron deposition. Clinical evaluation included standardized motor and cognitive testing. Longitudinal follow-up MRI was available for a subset of participants (20 fMC, 13 HC).

**Results:** fMC exhibited significant striatal atrophy compared to HC, with volume reductions in the caudate (−19.4%), putamen (−18.6%), and pallidum (−28.6%) (all *p*<0.001), despite the absence of overt movement disorders or cognitive deficits, accompanied by relative cerebellar hypertrophy. In fMC striatal volumes decreased with increasing age (putamen: rho=-0.382, p=0.012; caudate: rho=-0.631, p<0.001) and virtual time from genetically determined estimated age at onset (putamen: rho=-0.406, p=0.008; caudate: rho=-0.660, p<0.001). Susceptibility-weighted imaging revealed increased iron deposition in the caudate in fMC, which increased with greater local atrophy (*rho*=0.718, *p*<0.001). No significant longitudinal progression was detected over follow-up intervals of up to 25.5 months.

**Discussion:** Asymptomatic female carriers of the XDP-associated *TAF1* variant exhibit significant striatal neurodegeneration and iron accumulation, indicating a dominant-negative effect despite X-linked inheritance. Future longitudinal studies should focus on non-motor features and integrate genetic, imaging, and clinical markers to improve risk stratification and identify the subset of female carriers most likely to benefit from future disease-modifying therapies.

## Introduction

In many X-linked diseases traditionally considered recessive, affected women are rare.^1^ When clinical manifestations occur, they typically exhibit a late-onset and/or a mild phenotype. However, in recent years, penetrance and altered expressivity among female carriers of X-linked diseases have gained increasing attention, highlighting the need for a deeper understanding of their pathogenic mechanisms and clinical relevance.

X-linked dystonia-parkinsonism (XDP) is a severe neurodegenerative movement disorder affecting predominantly men with adult-onset dystonia and ensuing parkinsonism. Due to a founder effect, the disorder is confined to individuals of Filipino ancestry.^2^ The causal variant is an insertion of an SVA (short interspersed element (SINE)–variable number of tandem repeat (VNTR)–Alu)-type retrotransposon containing a polymorphic hexanucleotide repeat ((CCCTCT)n) within the *TAF1* gene on the X chromosome.^3^ The length of the repeat expansion modulates age at onset (AAO) and disease severity^4–6^, similar to other repeat expansion disorders.^7^ The existence of three additional genetic modifiers (tagged by three single-nucleotide polymorphisms, SNPs) influencing AAO was established in a genome-wide association study, and together with repeat length, the SNP constellation explains approximately 65% of AAO variability in men.^4^

Post-mortem examinations and structural quantitative MRI studies have identified pronounced striatal atrophy, particularly affecting the striosome-enriched subparts during the early dystonia-predominant disease stages in males^8,9^, as well as in the prodromal phase.^10^ In addition, increased iron deposition has been demonstrated in the anteromedial putamen of both manifesting and prodromal male *TAF1* variant carriers.^10,11^ Notably, prodromal male carriers examined about nine years before the predicted onset exhibited no clinical signs of dystonia or parkinsonism but could be distinguished from healthy controls by subtle alterations in gait and balance parameters using wearable sensors.^12^

To date, evidence in female XDP patients is limited to anecdotal reports and small case series.^13^ In line with other X-linked diseases, AAO in females is typically later than in male patients and is associated with generally milder and more variable phenotypes, including isolated parkinsonism, chorea, myoclonus, and atypical tremor, while dystonia tends to be less prominent.^14^

There are three proposed mechanisms that may explain disease penetrance in women^15^ all of which have also been described in XDP: (1) Homozygosity: A female homozygous carrier has been reported to exhibit a mild choreatic phenotype with late onset.^14^ (2) (Atypical) Turner syndrome: One female XDP patient with atypical (mosaic) Turner syndrome presenting with both, dystonia and parkinsonism, and a comparatively severe phenotype^16^ (3) Skewed X-chromosome inactivation (XCI): While XCI normally equalizes gene dosage between sexes and is usually balanced, it can become skewed, resulting in predominant expression of the mutated gene in most cells. An extremely skewed XCI pattern (98%:2%) was reported in a female XDP patient with isolated non-levodopa-responded Parkinsonism and typical XDP features on brain MRI.^17^

The possibility of skewed XCI is likely the most common of the above-mentioned mechanisms. In other X-linked diseases, such as Duchenne/Becker dystrophy and chronic granulomatous disease, skewed XCI has been shown to correlate with penetrance and expressivity in female mutation carriers.^18–20^

In this study, we investigated female carriers of the *TAF1* pathogenic variant causing XDP (fMC) without an overt movement disorder and healthy female controls (HC) using a multimodal approach including quantitative structural MRI and a deep clinical and genetic characterization. This study aimed to (1) identify subtle hyperkinetic or hypokinetic signs on clinical examination that remain unrecognized by the participants; (2) demonstrate striatal and pallidal neurodegeneration in fMC in the absence of an overt clinical manifestation, analogous to that observed in male prodromal mutation carriers; (3) detect incipient iron deposition in the anteromedial putamen, comparable to findings reported in male carriers; (4) examine associations between structural brain alterations and genetic markers, including the calculation of the estimated age at onset (eAAO), as established in male carriers^4^; and (5) assess longitudinal progression of basal ganglia degeneration in a subset of participants who underwent follow-up examinations after 18 or 25.5 months.

## Materials and methods

### Recruitment and Participants

The investigations took place at the Makati Medical Center, Manila, Philippines in May 2022, November 2023 and July 2024. Study information materials were provided in English and Tagalog and explained by a native Tagalog speaker. Before participation, written informed consent was given by all participants. The study was conducted according to the ethical standards of the revised version of the Declaration of Helsinki and approved by the local ethics committee of the Makati Medical Center.

One hundred three female relatives of male XDP patients were asked to participate in an extensive multimodal study that included clinical assessment, genetic analyses, balance and gait measurements, electroencephalography with event-related potentials, oculomotor recordings, transcranial sonography, and MR imaging. Given the inheritance pattern, an approximately 50% ratio of fMC to HC was anticipated. Genetic counselling was offered to all participants outside the study framework. All participants were advised in advance and agreed that they would not be informed of genetic or clinical testing results collected during the study. Since genetic testing was conducted only after data acquisition, all investigations were performed in a double-blind manner.

### Clinical and Genetic Assessment

Genetic testing revealed that 42 participants (age: mean (M) 53.7 years, standard deviation (SD) 12.82) were heterozygous for the disease-causing variant in the *TAF1* gene (fMC) whereas 61 participants (age: M 45.5 years, SD 12.10) carried two wildtype alleles (HC). Follow-up MRI examinations were done in 16 fMC and 12 HC after 18 months and in four fMC and two HC after 25.5 months.

fMC were significantly older than HC (t=-3.3, *p*=0.002), thus, age was included as a confounding variable in the subsequent analyses. Other genetic abnormalities, i.e., Turner syndrome and *TAF1* variant homozygosity, were excluded. In analogy to male hemizygous carriers, the repeat length of the hexanucleotide expansion (in the blood) and the three additional SNPs associated with AAO were analyzed in fMC to estimate the eAAO (if they were men) and the virtual time from manifestation (vTM) (age at examination subtracted by eAAO).^4^ The degree of XCI skewing was determined using a standard methylation-sensitive assay based on HpaII digestion of the androgen receptor (AR) gene, exploiting a polymorphic CAG repeat adjacent to the HpaII restriction site in exon 1 to discriminate between alleles in female participants, as previously described.^21,22^ XCI ratios ranging from 0:100 to 30:70 were classified as imbalanced. In samples exhibiting XCI imbalance, we determined the direction of skewing by sequencing a cDNA region encompassing disease-specific change 3 (DSC3; chrX:71529785C>T, GRCh38), the only transcribed variant within the XDP-specific haplotype. A predominance of the mutant T signal at the DSC3 position was interpreted as preferential expression of the XDP-associated allele carrying the SVA insertion, whereas a predominance of the reference C signal indicated preferential expression of the wild-type allele.

Whole-blood RNA was collected in PAXgene Blood RNA tubes (PreAnalytiX, Qiagen/BD), isolated according to the manufacturer’s instructions, and reverse-transcribed into complementary DNA (cDNA) using a commercially available kit (Fermentas, Thermo Fisher Scientific). Sanger sequencing of cDNA amplicons was performed on an ABI 3500XL Genetic Analyzer (Applied Biosystems).

The neurological examination was standardized, video-recorded, and performed by movement disorder experts (JU, NB). The dystonia and parkinsonism subscores of the Movement Disorder Society of the Philippines-XDP rating scale (MDSP-XDP Rating Scale I/II), the motor score of the Burke–Fahn–Marsden–Dystonia Rating Scale (BFMDRS), and the Unified Parkinson Disease Rating Scale: Part III: Evaluation of Motor Function (MDS-UPDRS-III) were used to evaluate motor signs. Cognitive functions were assessed with the Filipino version of the Montreal Cognitive Assessment (MoCA-P), the Frontal Assessment Battery (FAB), and the Trail-Making Test (TMT). Depressive symptoms and anxiety were investigated using the Filipino version of the Hospital Anxiety and Depression Scale (HADS).

### MRI Acquisition

T1-weighted and Susceptibility-weighted (SW) images were acquired on a 1.5T Magnetom Aera, Syngo MR D13 (Siemens Healthcare, Erlangen, Germany) equipped with a 20-channel head coil. A three-dimensional magnetization-prepared rapid acquisition echo gradient-echo sequence (sagittal orientation, 144 slices without gaps, 1×1×1 mm^3^ voxel size, field of view 256×256×144 mm^3^, flip angle 15°, repetition time 1900 milliseconds, echo time 2.66 milliseconds) was used for the T1-weighted images. The acquisition of the gradient-echo SW images was performed with the following settings: transverse orientation, 36 slices without gaps, voxel size 1×1×4 mm^3^, field of view 192×256×144 mm^3^, flip angle 15°, repetition time 49 milliseconds, echo time 40 milliseconds.

All images were inspected for movement artifacts and structural abnormalities (HH). Scans of one fMC and one HC had to be disregarded, resulting in 101 analyzable subjects for the cross-sectional analysis (41 fMC, 60 HC) and 33 subjects for the longitudinal analysis (20 fMC, 13 HC).

### Voxel-based Morphometry

Voxel-based morphometry (VBM) is a standard tool for assessing structural changes by using signal intensity to estimate the local amount of specific tissue (e.g., grey or white matter).^23^ The CAT toolbox (A Computational Anatomy Toolbox for SPM, Version 12.6) as an extension to Statistical Parametric Mapping 12 (SPM12) implemented in MATLAB R2017b

(MathWorks, Natick, Mass., USA) was used for preprocessing and analysis. The details have been described before.^10^ Because traditional absolute masking at a threshold of 0.1 would have blanked out two-thirds of the pallidum, this grey matter mask was expanded by an atlas mask of the pallidum (provided by Neuromorphometrics, Inc.) under academic subscription (originating from the OASIS project) with ImCalc. Independent t-tests with total intracranial volume, age, and timepoint of scanning as covariates were performed to compare fMC to HC at baseline.

For the longitudinal analysis, the CAT toolbox was used with default settings for preprocessing (except for the East Asian International Consortium of Brain Mapping (ICBM) template for affine regularization).^24^ Subsequently, an F test for interaction of time and group was performed.

Voxels exceeding the statistical threshold of p < 0.05 after cluster-wise family-wise error (FWEc) correction for multiple comparisons (initial threshold p< 0.001) were considered significant. Peak voxels were identified and labelled according to a maximum probability atlas provided by Neuromorphometrics.

Mean signal intensities were extracted from the significant clusters (fslstats, FSL utils, FMRIB Software Library v5.0, created by the Analysis Group, FMRIB, Oxford, UK) and implemented into SPSS 29.0 (IBM Corp, Armonk, NY) for further analysis and correlation with clinical and genetic data.

### Subcortical Volumetry

For the calculation of subcortical volumes, the standard protocol of the CAT toolbox was followed.^24^ The data were implemented into SPSS 29.0 for further analysis.

### Susceptibility-Weighted Imaging

SW imaging is a well-established MR imaging technique for clinical and research purposes among others to detect iron depositions.^25^ The SWI software provided by Siemens (based on the method first described by Haacke et al.^26^) automatically performed postprocessing of the phase and magnitude images to produce SW maps. Using SPM 12 in Matlab, SW images were brought to MNI space. The details have also been described before.^10^ Independent t-tests with age and timepoint of scanning as covariates and cerebrospinal fluid intensity as global value were performed to compare fMC to HC. In a whole-brain approach, voxels exceeding the statistical threshold of p < 0.05 after peak-wise FWE correction for multiple comparisons were considered significant. Peak-wise FWE correction was chosen instead of cluster-correction as the size of the expected cluster as well as the size of the structure in which the cluster was expected in combination with a relatively large voxel-size (1×1×4 mm) is too small to be detected in cluster-wise correction. For example, in prodromal male carriers, altered susceptibility could be demonstrated in two very small clusters in the putamen bilaterally (k= 17 and 21).^10^

Peak voxels were identified and labelled, and mean signal intensities were extracted from the significant clusters as described above.

### Statistical Analysis and Figure Creation

Statistical analysis was performed in SPSS 29.0. Kolmogorov-Smirnov tests were used to test for normal distribution. For non-normally distributed data, non-parametric tests were used (Mann-Whitney U-Test). T-tests and age-corrected analyses of covariance (ANCOVA) were conducted for normally distributed data. For correlation analyses, Spearman’s rho was applied. Outliers [as defined by Grubbs’ test (Grubbs, 1969)] were eliminated to prevent overestimated effects by single subjects.

GraphPad Prism (version 10.6.0 for macOS, GraphPad Software, La Jolla California USA, www.graphpad.com) was used for figure creation. Clusters were visualized on an averaged T1-weighted MRI scan of all participants with MRIcroGL (Version 26.0 for MacOs).

### Data availability

The data that support the findings of this study are available from the corresponding author, upon reasonable request.

## Results

### Clinical and genetical Assessment

Four fMC and five HC exhibited subtle, non-clinically significant signs, including mild choreatic hyperkinesias, parkinsonism, or dystonia. There were no differences in clinical scores for dystonia and parkinsonism between fMC and HC (mean scores are shown in Table 1). Likewise, no group differences were observed in executive functions, global cognition, anxiety and depression between fMC and HC (all *p*>0.15; please see Table 1 for details). The eAAO in fMC varied from 26.6 to 51.9 years resulting in a large range of vTM (−22.2 to 35.2 years). Whereas parkinsonian signs (MDS-UPDRS III score) increased with age at examination (AAE) in healthy controls (rho=0.310, p=0.017) but not in fMC (rho=0.280, p=0.080), dystonia (BFMRS) increased with AAE (rho=0.614, p<0.001) and vTM (rho=0.671, p<0.001) in fMC but not in healthy controls (rho=0.180, p=0.169). The MoCA total score decreased with vTM but not significantly (rho=-0.305, p=0.059) and showed no decrease with AAE in either HC (p>0.2) or fMC (p=0.197). The FAB total score decreased with both AAE (rho=-0.452, p=0.004) and vTM (rho=-0.472, p=0.002) in fMC, but not in healthy controls (p>0.2).

**Table 1.**
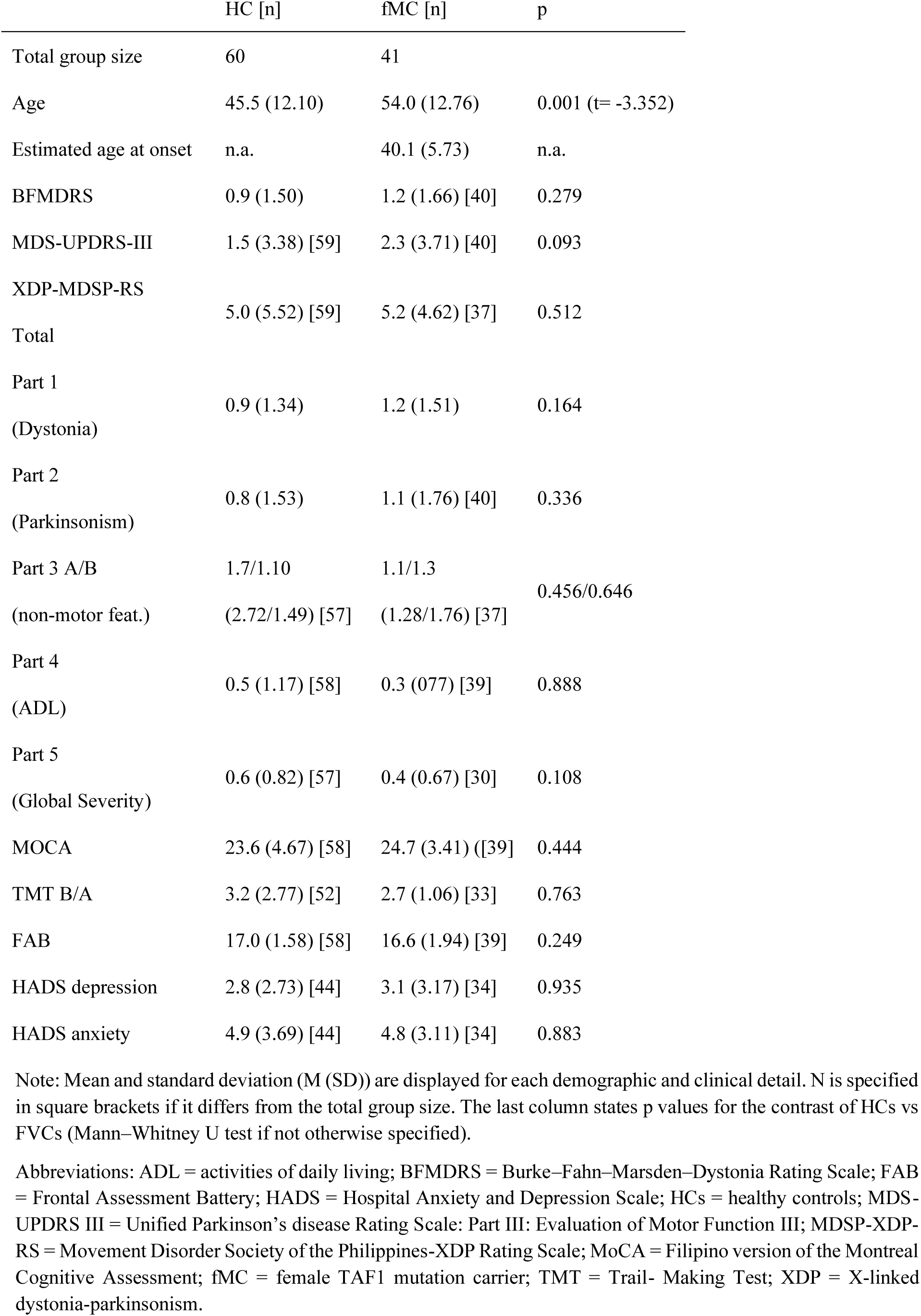
Overview of Demographic and Clinical Data.

XCI skewness was assessed in 28 fMC. Nine individuals exhibited skewed inactivation patterns (≥70:30), including two cases in which the mutant allele was predominantly expressed. No differences in basal ganglia volume were observed between individuals with random and skewed XCI patterns (MUT>WT and WT>MUT).

### T1-weighted imaging

Voxel-based morphometry at baseline revealed atrophy of the striatum and hypertrophy in the cerebellum in fMC compared to HC (Figure 1). Peak voxel coordinates are shown in Table 2.

**Figure 1:**
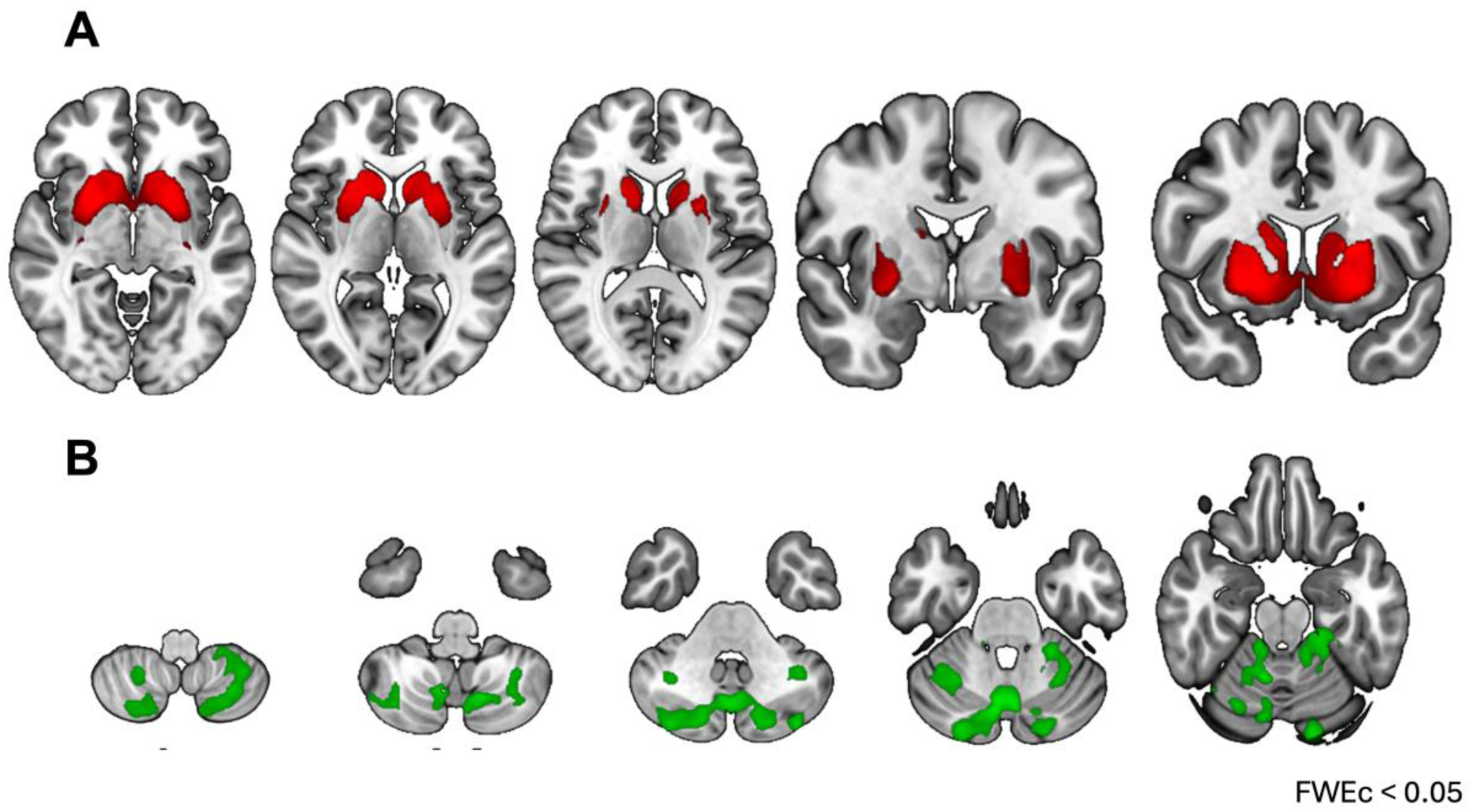
Voxel-based morphometry. (**A**) HC > fMC. (**B**) HC < fMC. Voxels exceeding the statistical threshold of 0.05 FWEc are color-coded. Color maps are superimposed on a T1-weighted 152-MNI template in neurological convention. HC: healthy controls, fMC: female mutation carrier. FWEc: cluster-wise family-wise-error-correction

**Table 2.** Significant Clusters of the Whole Brain VBM and SWI Analysis.

| Cluster Number | t-value | Size (k) | MNI Coordinates (mm) |  |  | Label |
| --- | --- | --- | --- | --- | --- | --- |
|  |  |  | X | Y | Z |  |
| Voxel-based morphometry |  |  |  |  |  |  |
| Controls > female mutation carriers |  |  |  |  |  |  |
| 1 | 9.23 | 29020 | 27 | 9 | -5 | Right putamen |
| Female mutation carriers > controls |  |  |  |  |  |  |
| 1 | 5.40 | 16555 | -20 | -89 | -19 | Left occipital fusiform gyrus<br>/ left cerebellum exterior |
| 2 | 3.98 | 2603 | -29 | -51 | -33 | Left cerebellum exterior |
| Susceptibility-weighted imaging |  |  |  |  |  |  |
| Controls > female mutation carriers |  |  |  |  |  |  |
| 1 | 5.14 | 18 | 11 | 10 | 10 | Right Caudate |

Note: The table depicts significant clusters of the whole brain VBM and SWI analysis with t-values, size of the cluster, MNI coordinates of the peak voxel, and labelling according to a maximum probability atlas provided by Neuromorphometrics, Inc. Please note that lower susceptibility values indicate higher susceptibility (i.e., higher tissue iron content).
Abbreviations: MNI= Montreal Neurological Institute; SWI= susceptibility-weighted imaging; VBM= voxel-based morphometry.

Accordingly, volumes of the caudate (*F*(1,98)=78.5, −19.4%), putamen (*F*(1,99)=55.1, - 18.6%), and pallidum (*F*(1,99)=18.6, −28.6%) were reduced in fMC compared to HC (all *p*<0.001, Figure 2). Caudate and putamen volumes decreased with increasing vTM in fMC

**Figure 2:**
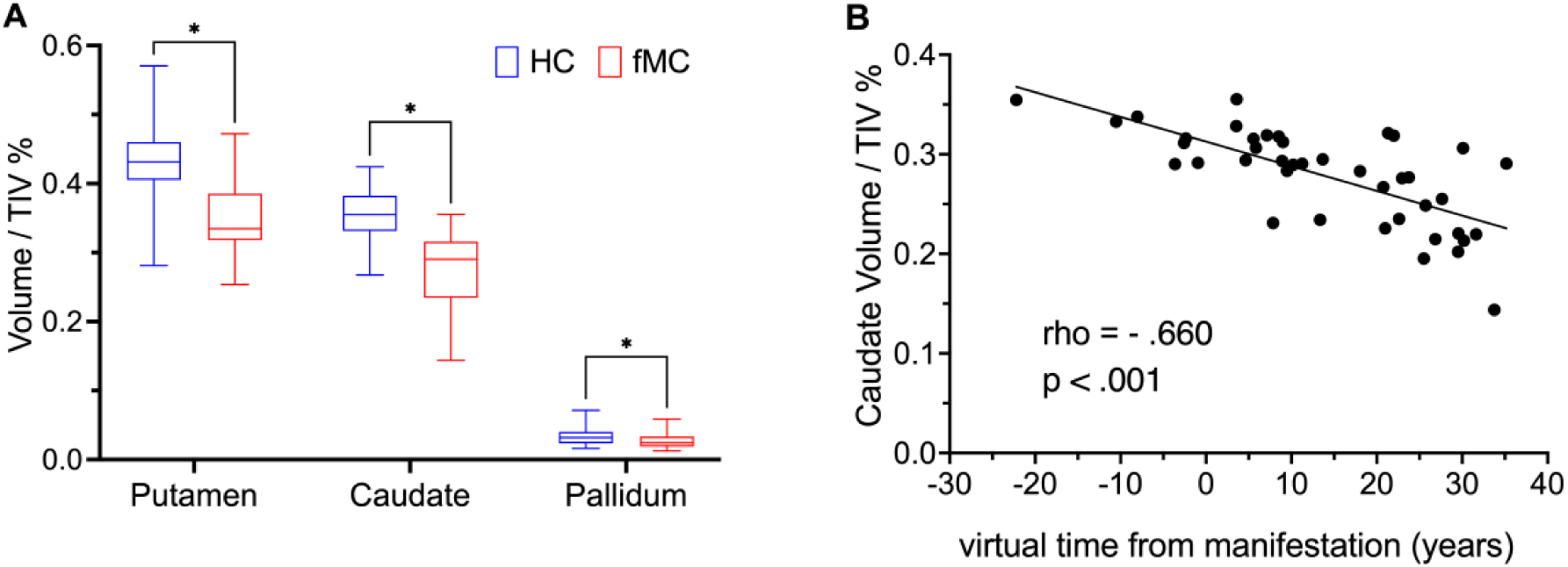
Subcortical volumetry. (**A**) Boxplots with whiskers represent mean and min. to max. values. Asterisks indicate a significant difference with p < 0.001. (**B**) Correlation with virtual time from manifestation. HC: healthy controls, fMC: female mutation carrier, TIV: total intracranial volume.

(putamen: rho=-0.406, p=0.008; caudate: rho=-0.660, p<0.001; Figure 3) and with increasing AAE (putamen: rho=-0.382, p=0.012; caudate: rho=-0.631, p<0.001). In contrast, pallidal volume showed a weak increase with increasing vTM (rho=0.319, p=0.039) and AAE (rho=0.366, p=0.017). In HC, only putaminal volume showed a weak decrease with increasing AAE (rho=-0.277, p=0.032).

**Figure 3:**
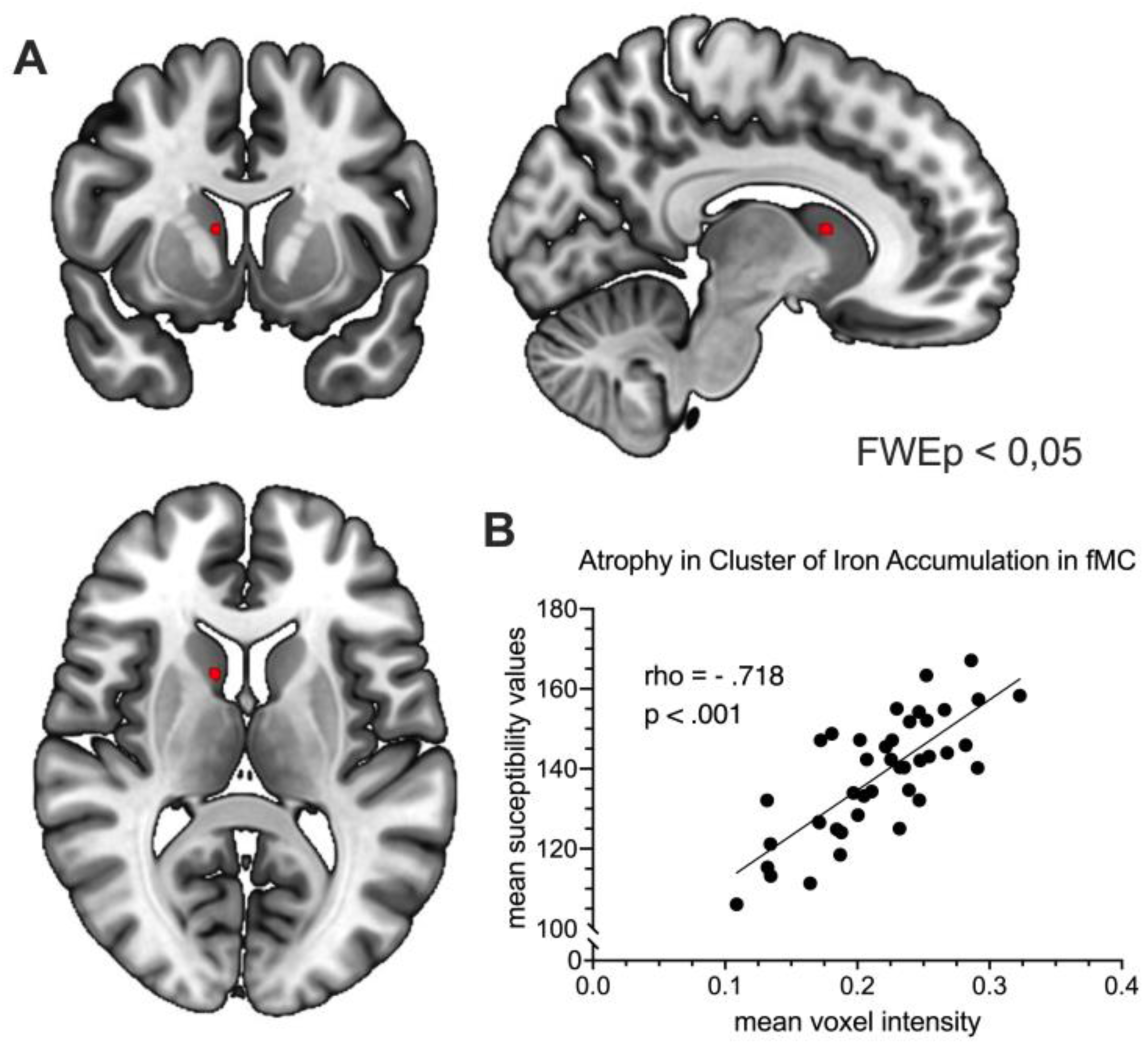
Susceptibility-weighted Imaging. (**A**) HC > fMC. Voxels exceeding the statistical threshold of 0.05 FWEp are color-coded. Color maps are superimposed on a T1-weighted 152-MNI template in neurological convention. Lower values indicate higher susceptibility (i.e., an increased iron content). (**B**) Correlation of grey matter atrophy and iron deposition. HC: healthy controls, fMC: female variant carrier. FWEp: peak-wise family-wise-error-correction

### Longitudinal T1-weighted imaging

In the longitudinal VBM analysis 21 months after baseline on average, the contrast of time and group showed no significant results. Likewise, there was no significant interaction of time and group in the subcortical volumetry of the pallidum, putamen and caudate nucleus. Individual trajectories of the caudate volume are shown in eFigure 1.

### Susceptibility-weighted imaging

SW imaging revealed increased iron accumulation in the caudate head in fMC compared to HC (Figure 3 and Table 2), whereas the reverse contrast yielded no significant clusters.

Extracted mean SW values of the significant caudate clusters correlated with mean voxel intensity in this region, indicating that iron deposition increased with greater atrophy in fMC (*rho*=0.718, *p*<0.001) but not in HC (*rho*=0.181, *p*=0.166).

## Discussion

We demonstrate that female carriers of the XDP-associated *TAF1* variant exhibit marked striatal neurodegeneration in the absence of overt clinical manifestations. Specifically, female variant carriers (fMC) showed significant atrophy of the caudate nucleus, putamen, and pallidum, accompanied by increased iron deposition in the caudate nucleus. These findings extend prior work in male patients and male prodromal carriers and indicate that XDP-related neurodegeneration is not confined to hemizygous males but is also present in heterozygous females at a subclinical stage.

The spatial pattern of atrophy observed in fMC is highly consistent with established neuropathological and imaging findings in XDP, which demonstrate a pronounced vulnerability of the striatum.^8,10,11^ In particular, early degeneration of striosome-rich compartments has been proposed to disrupt limbic–motor integration within basal ganglia circuits, thereby contributing to disease expression. ^9,27^ The magnitude of striatal volume loss in the present cohort suggests that these processes are already well advanced despite the absence of clinically detectable motor features, pointing to a substantial degree of clinicoradiological dissociation. This is consistent with evidence from other neurodegenerative disorders demonstrating that structural brain changes can precede clinical manifestation by many years^28–30^, likely reflecting compensatory network mechanisms and functional reserve. This marked dissociation between structural pathology and clinical phenotype in fMC also raises important questions regarding disease penetrance and compensation. However, more subtle phenotypes involving frontostriatal–limbic and cognitive–executive functions, behavioral changes, and oculomotor abnormalities were not investigated within the imaging-focused framework of the present study and should be addressed in future work, including an assessment of their clinical relevance. These aspects have been characterized using a complementary multimodal approach in our cohort and are the focus of separate analyses examining their relationship to the extent of neurodegenerative changes and to genetic factors.

One plausible mechanism underlying neural reserve is the partial preservation of functional *TAF1* expression through random XCI, which may buffer the impact of the mutant allele. At the same time, skewed XCI is a well-established determinant of phenotypic variability in X-linked disorders and likely contributes to the spectrum of subclinical and manifest disease observed in female carriers.^17–20^ In an exploratory analysis in a subgroup of fMC, basal ganglia volumes did not differ between individuals with random and skewed XCI patterns. However, the sample size is insufficient to draw robust conclusions. Moreover, XCI is known to be tissue-specific; thus, measurements obtained from lymphocytes may not reflect the XCI status in neuronal tissue. XCI patterns represent a critical target for future studies.

Another key finding of this study is the concomitant increase in cerebellar volume in fMC. Within the emerging framework of distributed motor network models, the cerebellum is increasingly recognized as functionally and anatomically interconnected with the basal ganglia via thalamic, brainstem and disynaptic pathways.^31,32^ In this context, cerebellar hypertrophy may reflect adaptive network-level reorganization in response to striatal dysfunction and degeneration. Similar observations in male mutation carriers, particularly in early stages^8^, suggest that such cerebellar involvement represents a general feature of XDP pathophysiology rather than a sex-specific phenomenon. Furthermore, in our previous work we demonstrated an upregulation of cerebellar energy metabolism, with increased levels of high-energy phosphate compounds in the cerebellum of fMCs and male XDP patients, providing additional evidence for a compensatory role of the cerebellum in carriers, independent of sex. In this small cohort of fMC (n=10), however, overall cerebellar grey matter volume was reduced compared with HC.^13^ This interpretation aligns with broader concepts of compensatory plasticity in neurodegenerative disorders but also non-degenerative dystonia, whereby structurally and functionally connected regions undergo adaptive changes to maintain network output.^33–35^ However, whether the observed cerebellar enlargement reflects true neuroplastic compensation, reactive gliosis, or methodological effects inherent to voxel-based morphometry remains unresolved and requires further investigation using longitudinal and multimodal approaches, particularly functional ones.

Striatal volumes decreased with increasing AAE and vTM in fMC, supporting a continuous and biologically meaningful trajectory of neurodegeneration. This observation suggests that genetic modifiers, including repeat length and associated SNPs, exert comparable effects across sexes, and reinforces the notion that female carriers can be positioned along a similar disease continuum as male patients.

Increased iron deposition in the caudate provides additional evidence of neurodegeneration in fMC. Brain iron accumulation has been implicated in oxidative stress and neuronal vulnerability^36–38^, and its association with regional atrophy in our cohort supports the convergence of these pathological processes. The apparent spatial shift compared to prior reports in male carriers may reflect differences in disease stage or sex-specific trajectories, although this warrants further clarification.

Longitudinal analyses did not reveal significant progression over the observation period. However, the association of baseline atrophy with age and eAAO supports ongoing and progressive neurodegeneration. The absence of detectable changes likely reflects limited statistical power, short follow-up duration, and heterogeneity in disease stage at follow-up. The slow progression observed in female carriers may explain why subtle parkinsonian features emerge only later in life. Whether symptomatic female carriers, potentially as a result of skewed XCI, contribute to the slight female predominance of parkinsonism reported in the Philippines remains to be determined as epidemiological evidence remains limited.^17,39^

Taken together, these findings challenge the traditional view of female carriers as unaffected and instead support a model in which heterozygous individuals occupy a subclinical or prodromal position within the XDP disease spectrum. The presence of significant striatal neurodegeneration suggests a dominant-negative effect of the mutant *TAF1* allele that is only partially mitigated by the second allele. Within this framework, skewed XCI likely represents a key modifier of disease expression.

More broadly, our results support a network-based model of XDP in which striatal degeneration is accompanied by adaptive changes in interconnected regions, particularly the cerebellum. These findings have important implications for disease modelling, biomarker development, and the design of future therapeutic strategies, including the potential inclusion of selected female carriers in early intervention approaches.

### Strengths and Limitations

This study provides the largest and most comprehensively characterized cohort of female *TAF1* variant carriers to date. The multimodal and double-blind design strengthens the validity of the findings and enables the detection of subclinical neurodegeneration. Importantly, integrating imaging and genetic markers enables positioning female carriers along the XDP disease continuum and provides novel insights into network-level mechanisms, including cerebellar involvement.

A key limitation of the present study is the rather small and, in particular, unequal sample size in the longitudinal analysis, which may have reduced statistical power to detect subtle structural changes over time. This constraint is inherent to studies involving asymptomatic mutation carriers in rare disorders, where recruitment is challenging and eligible individuals are limited. Moreover, participant inclusion was conducted in a double-blinded manner to ensure methodological rigor and to avoid selection bias. As a consequence, group sizes could not be predetermined but were determined probabilistically, further contributing to slight imbalances between subgroups. While these factors may have limited sensitivity to detect small longitudinal effects, they reflect the practical and ethical challenges of conducting prospective imaging studies in rare, genetically defined populations.

## Conclusions

This study demonstrates that female carriers of the XDP-associated *TAF1* variant exhibit significant striatal neurodegeneration despite the absence of overt clinical manifestation. These findings challenge the concept of female carriers as unaffected and instead position them within a subclinical disease spectrum. The study also demonstrates that known genetic modifiers exert a measurable biological effect, influencing the degree of neurodegeneration. The identification of concomitant cerebellar alterations further supports a network-based model of XDP. While the observed neurodegenerative changes appear to progress slowly over midlife and most female carriers are unlikely to develop overt disease, identifying the subset of individuals at greatest risk for future clinical conversion remains a key challenge. Future longitudinal studies should therefore focus on defining trajectories of structural and functional change, integrating genetic, imaging, and clinical markers—including executive, cognitive, and other non-motor features—to improve risk stratification and identify those female carriers who may ultimately benefit from disease-modifying interventions.

## Supporting information

eFigure 1

## Acknowledgments

We would like to sincerely thank the Department of Neuroscience, the Department of Pathology, Dr. Darwin Dasig, and the Board of Directors of the Makati Medical Center, Makati City, Metro Manila, and Hans Manalo and Jane Maranan for their assistance in organizing and executing the study. We thank Madita Grümmer for her work in study coordination.

During the preparation of this manuscript the author’s used ChatGPT (GPT-5.6 Luna, OpenAI) to improve readability and linguistic quality. After using this tool, the authors reviewed and edited the content as needed and take full responsibility for the content of the published article.

