## Supplementary material for "Preclinical Basal Ganglia Neurodegeneration in Female X-Linked Dystonia-Parkinsonism Carriers": eFigure 1

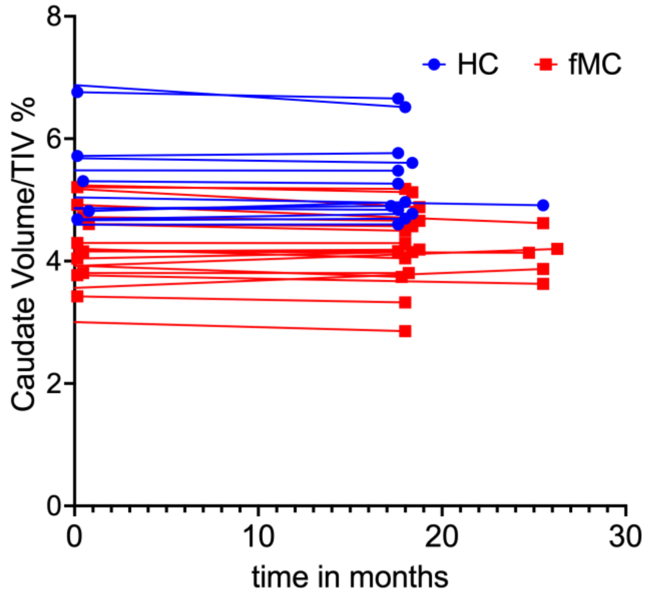


**eFigure 1: Longitudinal Volumetry of the Caudate Nucleus.**
Individual trajectories of caudate volume over follow-up intervals of 18 or 25.5 months.
HC: healthy controls, fMC: female mutation carrier, TIV: total intracranial volume.
